## Supplementary material for "Hydroxychloroquine/chloroquine for the treatment of hospitalized patients with COVID-19: An individual participant data meta-analysis": S1 Fig

**S1 Fig. Assessing Between-Study Heterogeneity**

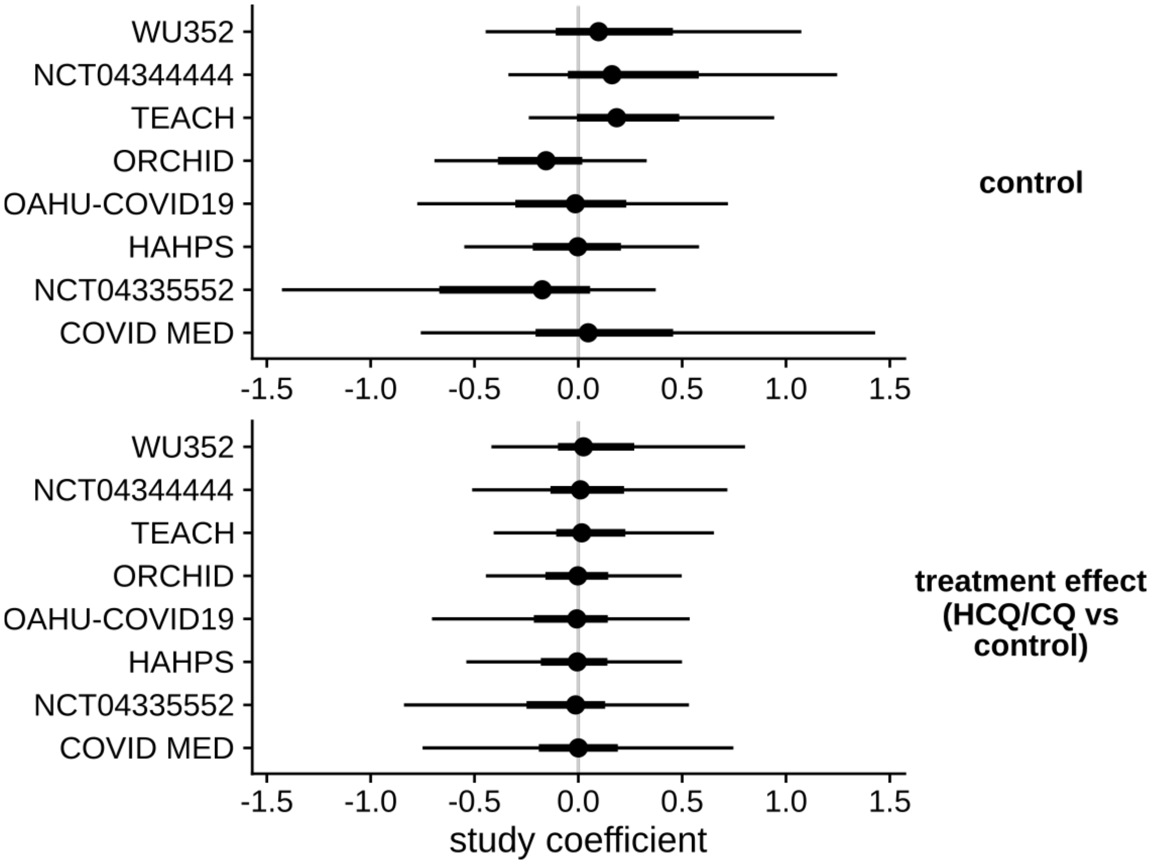


Estimated study coefficients from the primary outcome model, with 66% and 95% credible intervals (CrI) indicated. "Control" terms represent differences in predicted outcomes between sites under control, for individuals with similar baseline covariates, on the proportional log-odds scale. Treatment effect terms represent differences in predicted treatment effect between sites, again for individuals with similar baseline covariates. Positive coefficients represent, respectively, better ordinal scale outcomes under control and benefit of HCQ/CQ at days 28-35 post-enrollment. Estimated between-study standard deviations were 1.48 for the "Control" terms (95% CrI, 0.16-5.98; on the log-odds scale) and 0.87 for the treatment effects (95% CrI, 0.01-5.17). Tests of publication bias applied to unadjusted study-specific proportional ORs from the 6 studies with patients assigned to both HCQ/CQ and control yielded inconclusive results (Egger's test: p=0.10; Begg's test: p=0.57); these should be interpreted with caution given the small number of included studies.

HCQ/CQ indicates hydroxychloroquine or chloroquine.
