## Supplementary material for "Hydroxychloroquine/chloroquine for the treatment of hospitalized patients with COVID-19: An individual participant data meta-analysis": S1 Table

### S1 Table. Data Dictionary from Data Harmonization Spreadsheet

In addition to the variables below, we requested inclusion and exclusion criteria from each trial.

| Variable | Format | Description and comments |
|---|---|---|
| Patient ID | (Study-specific format) | |
| Other ID(s) | (Study-specific format) | |
| Treatment group | (Study-specific format) | |
| Enrollment date (year) | YYYY | Year of enrollment |
| Enrollment date (month) | Numeric (1-12) | Month of enrollment |
| Symptom onset date | Numeric (days) | Enrollment date - symptom onset date |
| Screening date | Numeric (days) | Enrollment date - screening date |
| Admission date | Numeric (days) | Enrollment date - admission date |
| Randomization date | Numeric (days) | Randomization date - enrollment date |
| Date of first dose | Numeric (days) | Date of first dose - date of enrollment (numeric; days) |
| Date of last dose | Numeric (days) | Date of last dose - date of enrollment |
| Ordinal outcome scale | Ordinal scale<br>1 = Death<br>2 = Hospitalized, on invasive mechanical ventilation or extracorporeal membrane oxygenation (ECMO)<br>3 = Hospitalized, on non-invasive ventilation or high-flow oxygen devices<br>4 = Hospitalized, requiring supplemental oxygen<br>5 = Hospitalized, not requiring supplemental oxygen<br>6 = Not hospitalized, limitation on activities<br>7 = Not hospitalized, no limitations on activities | Baseline ordinal scales as well as all available through day 30 post enrollment (Day 0, Day 7, Day 14, Day 28/30 are most important) |
| Date at which ordinal scale is measured | Numeric (days) | Date of ordinal outcome status – date of enrollment |
| Hospitalization length of stay | Numeric (days) | Duration in days between enrollment and day 28/30 post enrollment |
| Duration of mechanical ventilation | Numeric (days) | Duration in days between enrollment and day 28/30 post enrollment |
| All-cause mortality | 1 = Yes, 0 = No | Between enrollment and day 28/30 post enrollment |
| All-cause mortality – date of death | Numeric (days) | Between enrollment and day 28/30 post enrollment |
| Number of AEs | Numeric (count) | Between enrollment and day 28/30 post enrollment |
| Number of SAEs | Numeric (count) | Between enrollment and day 28/30 post enrollment |
| Number of AEs for QTC Prolongation | Numeric (count) | Between enrollment and day 28/30 post enrollment |

| Variable | Format | Description and comments |
| --- | --- | --- |
| Number of SAEs for QTC Prolongation | Numeric (count) | Between enrollment and day 28/30 post enrollment |
| Number of AEs for elevated liver function test | Numeric (count) | Between enrollment and day 28/30 post enrollment |
| Number of SAEs for elevated liver function test | Numeric (count) | Between enrollment and day 28/30 post enrollment |
| Number of AEs for arrhythmia or cardiac arrest | Numeric (count) | Between enrollment and day 28/30 post enrollment |
| Number of SAEs for arrhythmia or cardiac arrest | Numeric (count) | Between enrollment and day 28/30 post enrollment |
| Age | Numeric (years) | Age rounded *down* to beginning of 5-year bracket – e.g., 43 → 40; 39 → 35 |
| Sex | 1 = Male, 2 = Female | |
| Race | 1 = American Indian/Alaska Native;<br>2 = Asian;<br>3 = Black/African American;<br>4 = Native Hawaiian/Pacific Islander;<br>5 = White;<br>6 = Multiple;<br>7 = Other/declined;<br>8 = Unknown/unavailable | |
| Ethnicity | 0 = Not of Hispanic, Latinx, or Spanish origin<br>1 = Hispanic, Latinx, or Spanish origin<br>2 = Unknown | |
| BMI | numeric | |
| On mechanical ventilation at enrollment | 1 = Yes, 0 = No | |
| AIDS (do not include HIV-positive without AIDS criteria) | 1 = Yes, 0 = No | |
| Cerebrovascular disease | 1 = Yes, 0 = No | |
| A prior myocardial infarction | 1 = Yes, 0 = No | |
| Congestive heart failure | 1 = Yes, 0 = No | |
| Dementia | 1 = Yes, 0 = No | |
| COPD | 1 = Yes, 0 = No | |
| Asthma | 1 = Yes, 0 = No | |
| History of hypertension | 1 = Yes, 0 = No | |
| HIV positive (without AIDS) | 1 = Yes, 0 = No | |
| Solid tumor | 1 = Yes, 0 = No | |
| Liver disease | 1 = Yes, 0 = No | |
| Diabetes mellitus | 1 = Yes, 0 = No | |
| Cigarette or tobacco smoking | 1 = Yes, 0 = No | |
| Vaping | 1 = Yes, 0 = No | |
| Concurrent corticosteroid use | 1 = Yes, 0 = No | |
| Concurrent azithromycin use | 1 = Yes, 0 = No | |
