## Supplementary material for "Hydroxychloroquine/chloroquine for the treatment of hospitalized patients with COVID-19: An individual participant data meta-analysis": S2 Appendix

### S2 Appendix. Methods supplement: Description of the primary outcome model and estimands

**Model Description**

We fit a Bayesian proportional odds ordinal regression model for ordinal score measured between day 28 and day 35 post enrollment. For individuals with multiple measurements, the outcome was taken as the earliest measurement within the time window. Deaths recorded before the time window were carried forward. The following baseline covariates were included in the model: sex, age, number of comorbidities, body mass index, COVID ordinal scale). The coding and reference levels for the baseline variables are indicated in the table below.

| Covariate | Coding | Reference level | Binned version |
|---|---|---|---|
| Sex | female = +1/2, male = -1/2 | 0 (midpoint) | n/a |
| Age | (age in years – 60)/10 | 60 years old | <18, 18-29, 30-49, 50-69, 70-79, 80+ |
| Number of baseline comorbidities | raw count of the following baseline comorbidities | 0 | 0, 1, 2, 3, ≥4 |
| Body mass index | (BMI – 25)/5 | BMI of 25 | ≤20, 20-25, 25-30, 30-35, ≥35 |
| Baseline ordinal scale | indicators for levels 2-5, as well as (5 – the numeric score) | 5 = hospitalized, not requiring supplemental oxygen (the highest possible value for inpatients) | NA |

Missing baseline covariate data was imputed using multiple imputation via the R package mice (version 3.12) [1]. Treatment assignment and outcome were not used the imputation process. All posterior computations described below were pooled across the imputations. Individuals with missing outcome data were excluded from the model fitting; individuals with missing baseline data on a given covariate were excluded from the corresponding subgroup effect estimates.

Let $i$ index an individual patient. Each patient has a vector of baseline covariates $X_i$ and a treatment assignment $T_i$ (1 for hydroxychloroquine or chloroquine or 0 for control). Let the primary outcome for individual $i$ be denoted by $Y_i$ with levels $l$ = 1, …, 7. The proportional odds model takes the form:

$$logit\ P[Y_i \leq l\ |X_i, T_i] = \theta_l - \eta_i;\ l = 1, \ldots, 6$$

where $\theta_1, .., \theta_6$ are cutpoints that are common to all individuals (prior - ordered Student t prior with 3 degrees of freedom and scale parameter 2.5);

$$\eta_i = Z_i^T \beta_0 + \alpha_{0,study} + \delta_{0,NCOSS} + T_i\ (\tau + Z_i^T \beta_1 + \alpha_{1,study} + \delta_{1,NCOSS})$$

is the linear predictor; $Z_i$—a function of $X_i$—is a vector including sex, natural cubic splines (with 3 degrees of freedom) for age, BMI and number of comorbidities, and 5 minus ordinal score; $\beta_0$ and $\beta_1$ are fixed effects (prior - uniform); $\tau$ is a fixed effect (prior – uniform); $\delta_0$ and $\delta_1$ are independent normally distributed mean zero random effects (prior for standard deviation parameters — a half Student-t distribution with 3 degrees of freedom and scale parameter 10).

The model was fit using R, and the library “brms” (version 2.15) [2,3].

**Effect Estimates of Interest**

We produced two kinds of effect estimates:

**1. Standardized effect estimates**. These represent the effect of the treatment, averaged over the empirical distribution of individual-level covariates.

We estimated two effects: a *proportional odds ratio* and *risk difference for mortality*. We produced these estimates as follows.

Let $\pi_i^l(t)$ denote the predicted probability that an individual with covariates $X_i$ has outcome level $l$ under treatment $t$. Let $\pi^l(t) = \frac{1}{n}\sum_{i=1}^{n} \pi_i^l(t)$ be the predicted probability of outcome level $l$ under treatment $t$ in a population with the same covariate distribution as in our study.

*Proportional Odds:* For each iteration of the MCMC algorithm, our estimation of the standardized effect is tantamount to estimation by simulation with the following three repetitive steps: (1) draw a vector of covariates from the empirical distribution; (2) compute the predicted outcome probabilities under treatment and under control; and (3) use these predicted probabilities to draw outcomes under treatment and control (independently). Repeat steps (1) to (3) to generate a very large dataset and fit a proportional odds model with treatment indicator as the sole covariate. The resulting regression coefficient (i.e., log proportional odds ratio) is our standardized treatment effect on this iteration of the MCMC. In practice, for each iteration of the MCMC we fit a weighted proportional odds model with treatment as the sole covariates to a dataset with seven outcome levels crossed with two treatment levels; the weights for outcome level $l$ and treatment level $t$ are proportional to $\pi^l(t)$.

The associated estimand is the odds ratio from the closest fitting proportional odds model (with treatment as the sole covariate) to the true outcome probabilities under treatment and under control for a population with same distribution of covariates as in our pooled dataset.

*Risk Difference:* We utilize the procedure above with the exception that there is no need to fit the proportional odds model; we simply utilize the predicted probabilities of death under treatment and control and compute the difference. The estimand is the true risk difference under treatment versus control for a population with same distribution of covariates as in our pooled dataset.

*Subgroup effects:* We apply the above estimation procedure where we restrict the covariate distribution to the specific subgroup of interest. The estimands are subgroup specific.

**2. Conditional effect estimates.** For each level of a given covariate of interest and within iteration of the MCMC, we compute (1) predicted probabilities of mechanical ventilation or death under treatment and under control as well as the associated relative risk and (2) difference in the values of the linear predictor under treatment and under control, with all other covariates set to their reference values. In these computations, the quantities are marginalized over the study random effects. Posterior summaries of these quantities are plotted.

**Model Diagnostics and Sensitivity Analysis**

To assess the within-sample fit of our model, we compared the observed outcome data with draws from the posterior predictive distribution and examined Dunn-Smyth randomized quantile residuals [4].

To assess the sensitivity of our conclusions to modeling choices, we:

- repeated the analysis with weakly informative $N(0, 5^2)$ priors on the fixed effect coefficients, and more conservative half Student-*t* (df = 3, scale = 5) priors on the group-level standard deviations,
- explored the impact of adding to the model the additional variables (1) randomization to treatment with azithromycin and (2) time between symptom onset and enrollment, and
- (post-hoc) fit a version of the model without individual-level treatment interaction terms.

To compare the model fits, we recomputed the primary outcome and mortality estimates as well as estimated leave-one-out predictive (log) densities (LOO-ELPD) using the R package "loo" [5,6]. The results are as follows:

| | Relative LOO-ELPD | Standard error |
|---|---|---|
| Prespecified model minus interaction terms | 0.0 | 0.0 |
| Prespecified model with weakly informative priors | -4.2 | 4.0 |
| Prespecified model | -6.1 | 4.6 |
| Prespecified model with azithromycin and days since symptom onset terms | -9.6 | 4.8 |
| Prespecified model fit only to the ORCHID data, and without site effects | -54.2 | 11.6 |

These results indicate that the model without interaction provides the lowest cross-validated error. See main manuscript for our interpretation of these findings.
