## Supplementary material for "Hydroxychloroquine/chloroquine for the treatment of hospitalized patients with COVID-19: An individual participant data meta-analysis": S2 Fig

### S2 Fig. Conditional Covariate Effects

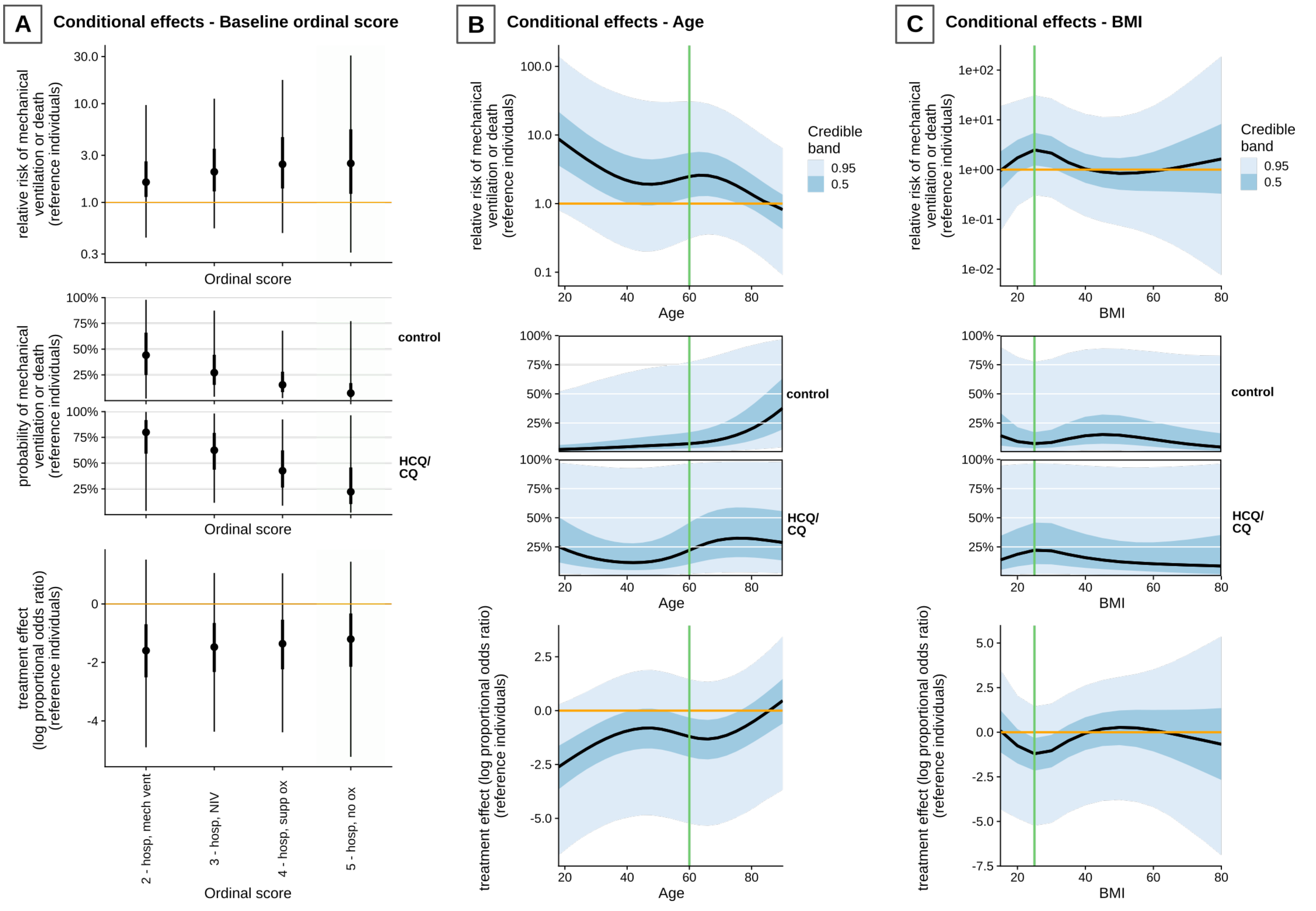

***S2 Fig (continued)***

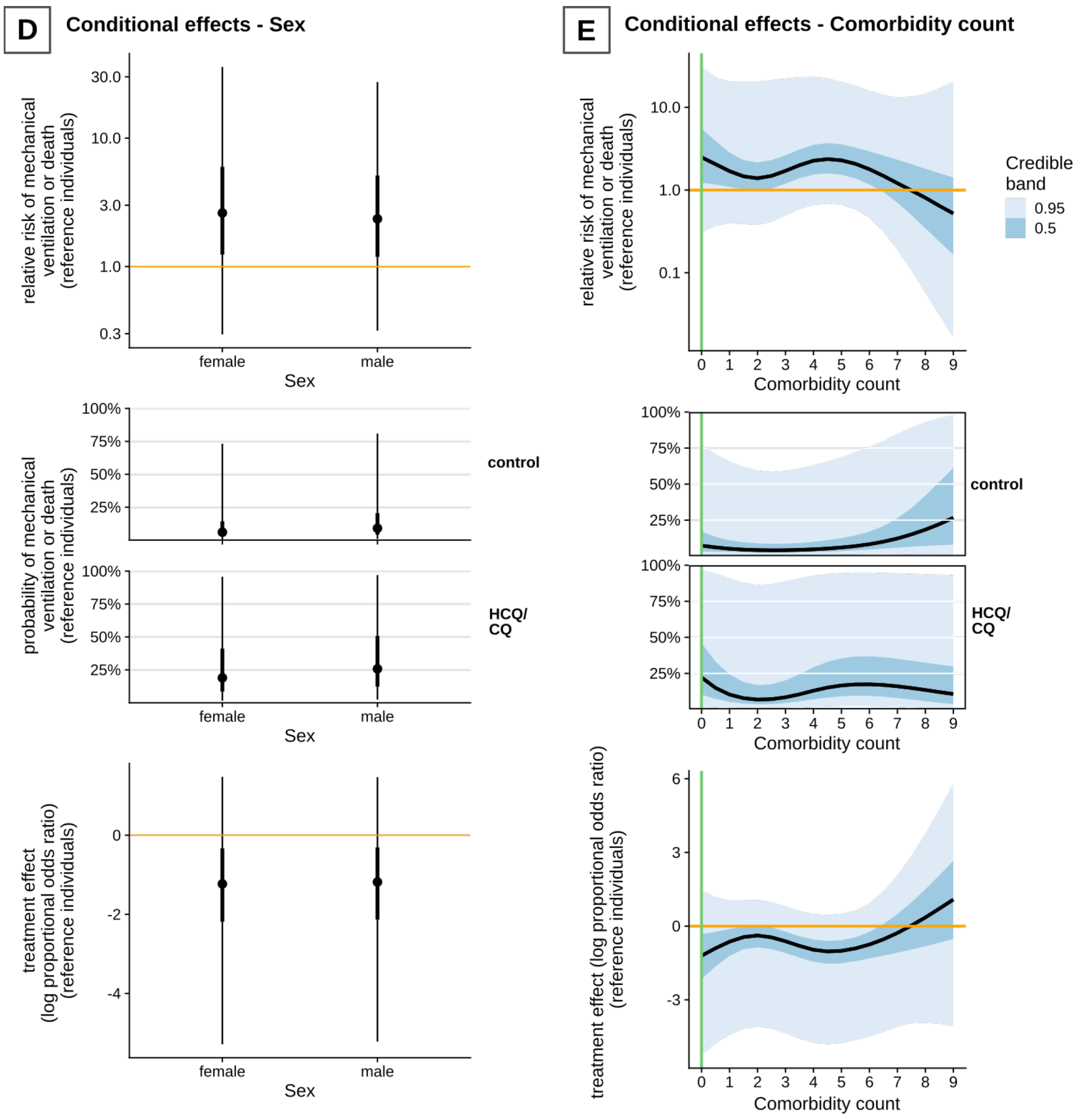


Conditional effects from the Bayesian proportional odds model. Shown are (1) the relative risk of mechanical ventilation or death at day 28-35; (2) the estimated probabilities of mechanical ventilation or death at day 28-35 under control and HCQ/CQ; and (3) the log proportional odds ratio comparing HCQ/CQ and control. Each of these effects are shown for reference individuals with the following covariate values: age 60, BMI 25, no baseline comorbidities, baseline ordinal score of 5, and sex coefficient set between male and female values. Curves for continuous covariates are accompanied by 50% and 95% credible bands; intervals for discrete covariates are accompanied by 66% and 95% credible intervals.

BMI indicates body mass index; HCQ/CQ, hydroxychloroquine or chloroquine.
