## Supplementary material for "Hydroxychloroquine/chloroquine for the treatment of hospitalized patients with COVID-19: An individual participant data meta-analysis": S2 Table

### S2 Table. Risk of Bias Assessment

| Trials | Bias arising from randomization process | Bias due to deviations from intended intervention | Bias due to missing outcome data | Bias in measurement of outcome | Overall risk of bias |
|---|---|---|---|---|---|
| ORCHID | Low | Low | Low | Low | Low |
| WU352 | Low | Low | Low | Some concerns | Some concerns |
| NCT04335552 (Duke) | Low | Low | Low | Some concerns | Some concerns |
| TEACH | Low | Low | Some concerns | Low | Some concerns |
| COVID MED | Low | Low | Low | Low | Low |
| HAHPS | Some concerns | Low | Low | Some concerns | Some concerns |
| NCT04344444 (University Medical Center New Orleans) | Some concerns | Some concerns | Low | Some concerns | Some concerns |
| OAHU-COVID19 | Low | Some concerns | Low | Some concerns | Some concerns |

We utilized the Cochrane Risk of Bias 2 (RoB 2) tool to rate specific risk of bias domains and "overall risk of bias." We considered the following domains of bias, using trial protocols, IPD, and other information provided by investigators: (1) bias arising from the randomization process (methods used to generate and conceal the allocation sequence), (2) bias due to deviations from intended interventions (whether participants and health professionals were masked to assigned intervention and methods used to ensure that participants received allocated intervention), (3) bias due to missing outcome data, and (4) bias in measurement of the outcome. Since we analyzed IPD, we excluded the fifth domain "risk of bias in selection of the reported result." We followed the recommended algorithms to reach an overall "risk of bias" assessment for each trial.

We assessed 6 studies at low risk of bias in the first domain of "Bias arising from the randomization process" (ORCHID, WU352, NCT04335552, TEACH, COVID MED, OAHU-COVID19); COVID MED was assessed as low risk because despite allowance for study arm shifting due to drug supply interruption, this never took place. Two studies (HAHPS, NCT04344444) did not have information on allocation concealment, and NCT04344444 had noticeably different sample sizes for each treatment group. All but two studies had low risk of bias for the second domain of "Bias due to deviations from the intended intervention"; NCT04344444 and OAHU-COVID19 elicited some concerns due to lack of information in the former and no masking in the latter. TEACH was the only study to elicit some concerns for "Bias due to missing outcome data" due to data missing for >20% of the study population. Finally, 5 studies scored "some concerns" for the domain of "Bias in the measurement of the outcome" (WU352, NCT04335552, HAHPS, NCT04344444, OAHU-COVID19). The primary reason for this rating was due to the outcome assessors being aware of the treatment each participant received; the rating was not assessed as "high risk of bias" because our primary outcome measurement is a hard endpoint for which most decision making is protocolized by objective oxygenation and ventilation respiratory status numbers. Our "Overall risk of bias" assessment was "low" for 2 and "some concerns" for 6 of the 8 studies.
