## Supplementary material for "Hydroxychloroquine/chloroquine for the treatment of hospitalized patients with COVID-19: An individual participant data meta-analysis": S3 Appendix

### S3 Appendix. Search Strategy in Detail

This study was initiated during extensive efforts by multiple organizations to promote collaborative research during the pandemic. Starting on April 30, 2020, a National Institutes of Health (NIH)-funded Trial Innovation Network team aiming to pool randomized clinical trial (RCT) data performed systematic outreach to the NIH Clinical and Translational Science Awards (CTSA) Program community. We connected with interested study teams via the COVID-19 Collaboration Platform website (http://covidcp.org) and additional directed contacts. Trialists were encouraged to upload protocols to the COVID-19 Collaboration Platform repository in an effort to promote collaboration and data aggregation and reduce duplication. We utilized this multi-pathway approach to assure the widest possible knowledge of our effort across established groups of likely investigators in the United States.

One site contacted by the COVID-19 Collaboration Platform, Bassett, had initiated a collaboration registry effort and performed systematic searches of ClinicalTrials.gov using search terms "COVID-19" and "hydroxychloroquine OR chloroquine," with a study status of "recruiting," on May 9, 2020, yielding 9 COVID-19 HCQ/CQ RCTs, and again on May 21, 2020; the latter search and subsequent recruiting elicited an initial list of 19 RCTs (18 from ClinicalTrials.gov, 1 from personal communication). PIs from studies in the Bassett/COVID-19 Collaboration Platform list were invited to participate in an HCQ/CQ pooled analysis project; outreach was initiated by Bassett and subsequently completed by the Trial Innovation Network team. Although research community outreach was an important component of our process, our study selection was dictated by the ClinicalTrials.gov searches. Of the 19 studies, we selected 8 after excluding 2 that declined participation or did not respond; 3 with ineligible trial designs including outpatient and prophylaxis studies; 2 with no enrollment; 1 not registered on ClinicalTrials.gov (identified via personal communication); and 3 with sites located outside the US (e.g., RECOVERY and DisCoVeRy, part of SOLIDARITY). (We sought to avoid cumbersome international data sharing regulatory delays such as the General Data Protection Regulation.) We were able to include 93.3% of patients from the targeted list of US studies. Excluding data from the international trials in our original list, on the other hand, led to the exclusion of a large number of patients: 6,569 in total.

Simultaneously, the Trial Innovation Network team completed a systematic search of ClinicalTrials.gov using search terms "COVID-19 OR SARS-CoV-2," with the country restricted to United States, on June 2, 2020, yielding 207 studies. That list was filtered to CTSA or CTSA affiliate sites in part because of consistent data collection at these sites, yielding 103 studies. The team surveyed this CTSA consortium; 26 expressed interest in collaboration. These searches and outreach activities, in addition to review of the studies' key variables, resulted in identifying 13 HCQ/CQ studies. A consensus investigator meeting made the decision to focus on inpatient trials. The rationale was that despite a substantial event rate occurring in inpatient trials, many were experiencing recruitment difficulty. Conversely, the outpatient trials, with lower event rates, had robust recruitment and desire to complete recruitment prior to pooling. Of the 13 trials, 10 were excluded: 4 with prophylactic trial designs, 2 outpatient trials, and 4 with no enrollment (some were excluded for more than one reason). The outpatient studies were redirected to an outpatient pooling effort (Gates Foundation). Five of the Trial Innovation Network's list of 13 trials overlapped with the Bassett/COVID-19 Collaboration Platform list, and 3 of those were included in our analysis (ORCHID, WU352, COVID MED).

The list developed by Bassett and the COVID-19 Collaboration Platform was the primary driver for study inclusion/exclusion in our pooled analysis, with augmentation and refinement by the Trial Innovation Network's outreach and search efforts. None of the selected studies had been published prior to selection.

#### Summary of Search Strategies

| | **COVID-19 Collaboration Platform/Bassett** | **Trial Innovation Network** |
|---|---|---|
| **General search description** | | |
| Study registry searched | ClinicalTrials.gov | ClinicalTrials.gov |
| General search strategy | Keywords | Keywords |
| Search date(s) | May 9, 2020; May 21, 2020 | June 2, 2020 |
| **Search strategy used** | | |
| | COVID-19 AND (hydroxychloroquine OR chloroquine) | COVID-19 OR SARS-CoV-2 |
| | Status: Recruiting | Country: United States |

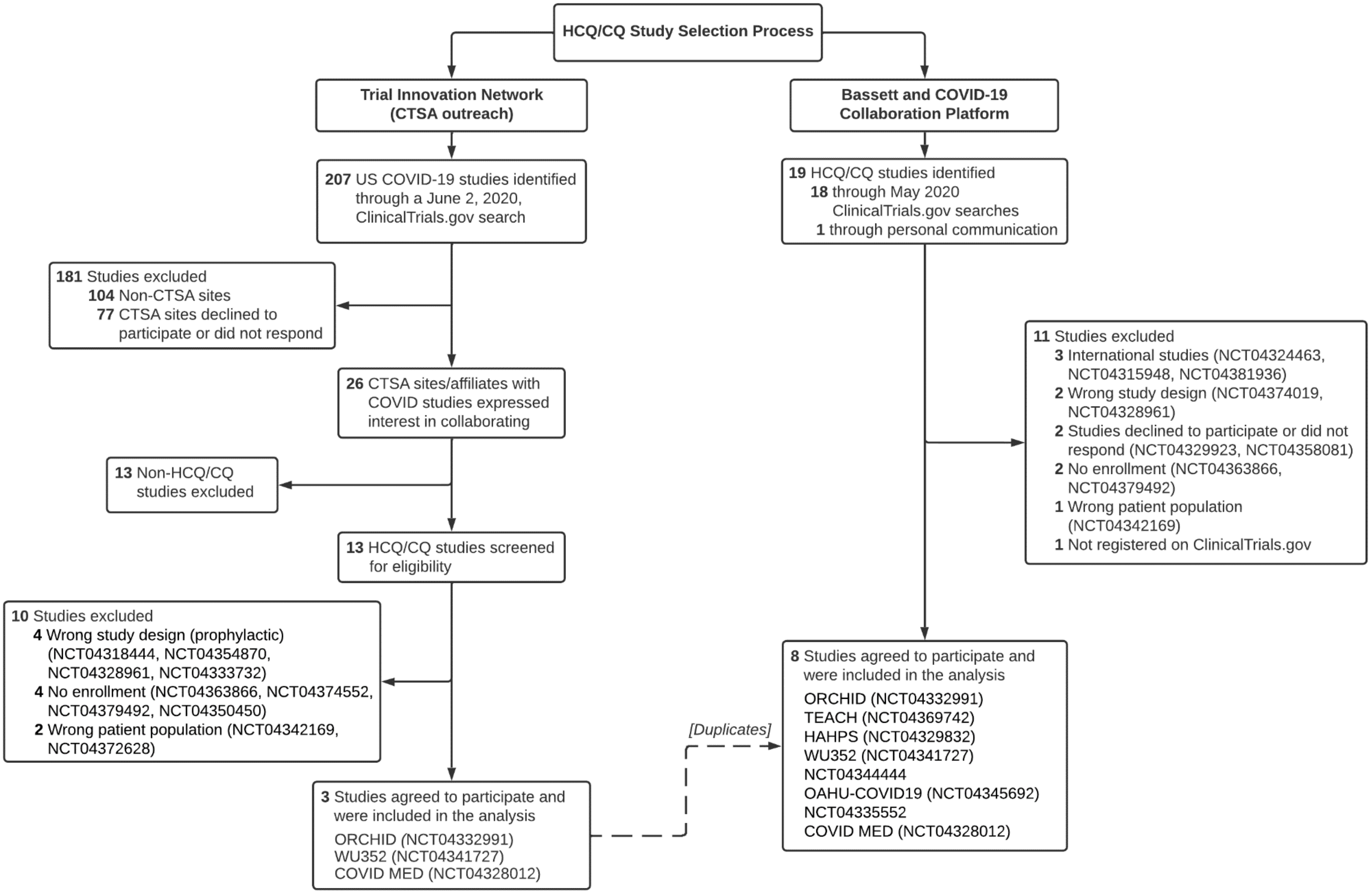


**Study Selection: Extended Version.** Two of the trials included in our analysis did not have study acronyms (only trial registration numbers). COVID MED indicates Comparison Of Therapeutics for Hospitalized Patients Infected With SARS-CoV-2 In a Pragmatic aDaptive randoMizED Clinical Trial During the COVID-19 Pandemic; CTSA, Clinical and Translational Science Awards; HAHPS, Hydroxychloroquine vs. Azithromycin for Hospitalized Patients With Suspected or Confirmed COVID-19; HCQ/CQ, hydroxychloroquine or chloroquine; OAHU-COVID19, A Randomized, Controlled Clinical Trial of the Safety and Efficacy of Hydroxychloroquine for the Treatment of COVID-19 in Hospitalized Patients; ORCHID, Outcomes Related to COVID-19 Treated With Hydroxychloroquine Among In-patients With Symptomatic Disease; TEACH, Treating COVID-19 With Hydroxychloroquine; WU352, Washington University 352: Open-label, Randomized Controlled Trial of Hydroxychloroquine Alone or Hydroxychloroquine Plus Azithromycin or Chloroquine Alone or Chloroquine Plus Azithromycin in the Treatment of SARS CoV-2 Infection.
