## Supplementary material for "Hydroxychloroquine/chloroquine for the treatment of hospitalized patients with COVID-19: An individual participant data meta-analysis": S3 Fig

**S3 Fig. Estimated Mortality Rate in Subgroups Under Both Control and HCQ/CQ**

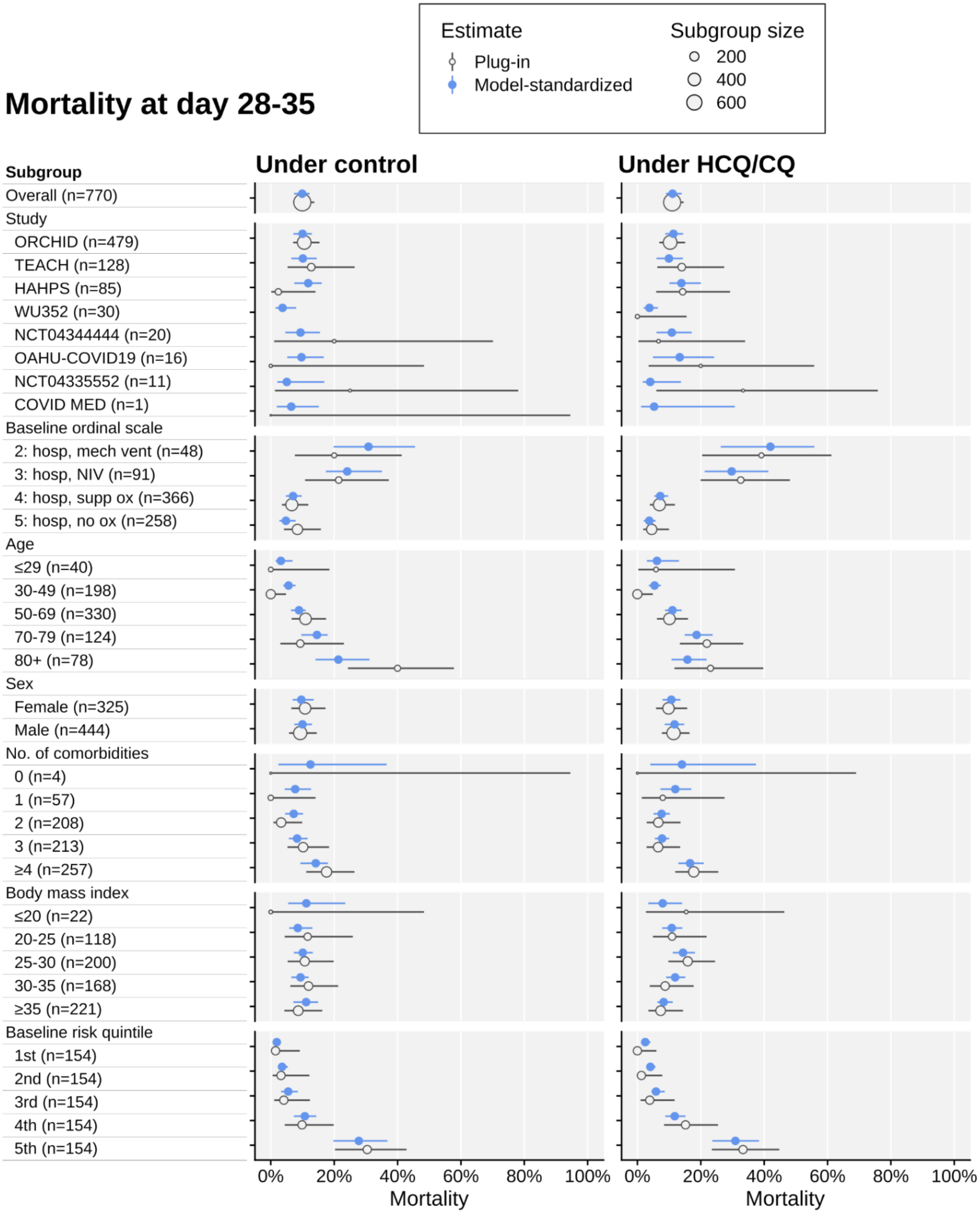


Shown are both plug-in estimates (based on the proportion of deaths in each subgroup) along with 95% CIs, and model-adjusted estimates with 95% credible intervals. The model used is the same as for the primary outcome analysis.

HCQ/CQ indicates hydroxychloroquine or chloroquine.
