## Supplementary material for "Hydroxychloroquine/chloroquine for the treatment of hospitalized patients with COVID-19: An individual participant data meta-analysis": S3 Table

### S3 Table. Changes to the Prespecified Statistical Analysis Plan

| When the change was made | Change | Reason |
|---|---|---|
| After examining baseline covariates and outcome missingness, but not outcome data themselves | We decided to extend the outcome definition from day 28-30 post-enrollment to day 28-35 post-enrollment. A patient's outcome was taken to be their earliest recorded ordinal score between day 28 and day 35 post-enrollment (inclusive). | Missingness in the outcome measurements. The change increased the number of patients with valid outcome values from 90.3% to 95.3%. |
| | We decided to use a simple count of those comorbidities without significant missingness in place of a weighted Charlson (or Charlson-like) comorbidity score. | Missingness in baseline comorbidity indicators; not having requested standard Charlson indicators from each site. |
| | We simplified the safety variables under consideration. | Extensive missingness in QTc and elevated LFTs AE/SAE results. |
| | We modified the form of our prespecified regression model for the primary outcome. | Following establishing the total sample size and simulating outcome data from the empirical distribution of baseline patient characteristics. |
| After examining/analyzing the outcome data | We decided to use superpopulation rather than finite sample standardized estimators of treatment effect for our primary outcome analysis. | For three reasons: (1) the uncertainty in the superpopulation estimator is more directly comparable to that of the maximum likelihood estimator; (2) the finite-sample estimator requires assumptions about the dependence between individual-level potential outcomes; and (3) our choice of assumption—to treat the potential outcomes as independent—potentially made the associated uncertainty intervals misleadingly narrow. |
| | We set as missing BMIs less than 10 and greater than 70; for the outcome analysis, these were imputed in the same step as the other baseline covariates using multiple imputation. | Extremeness of these values. |
| | We decided not to fit a category-specific ordinal model as a sensitivity analysis. | Time and effort; the reasonable within-sample fit of the simpler models. |

| When the change was made | Change | Reason |
|---|---|---|
| | We decided to de-emphasize our pre-specified conditional effect measure (relative risk of mechanical ventilation/ECMO or death). | The associated uncertainty intervals were extremely wide, perhaps due to the flexibility of our prespecified model. |
| | We decided to include model-standardized estimates of the risk difference for mortality, both overall and by subgroup. | This was considered informative and straightforward, given the model. Additionally, risk differences are considered a more interpretable measure of subgroup effects than odds ratios because of their collapsibility [1,2]. |
| | We decided to include a subgroup analysis based on quintiles of a baseline risk score. | Following recommendations of Kent et al [3]. |
| | We decided not to examine whether site × treatment interactions are associated with site-level covariates or individual-level covariates averaged within sites. | There was very little variation in the estimated site × treatment interactions. |
| | We replaced mortality at day 28-30 as a safety outcome with mortality at day 28-35 as a secondary outcome, and conducted an analysis of this parallel to that of our primary outcome. | We had prespecified all-cause mortality at or before day 28/30 as a safety outcome. However, we judged that an analysis of mortality parallel to that of our primary outcome would be clinically relevant. |
| | In the primary outcome and mortality analyses, we treated 6 extreme BMI values (<10 or >70) as missing. | We suspected that these values were mistaken or could bias our results, and were unable to definitively establish their accuracy. |
| | We added an exploratory post-hoc subgroup analysis based on time between symptom onset and enrollment. | Suggested in review. |
