## Supplementary material for "Hydroxychloroquine/chloroquine for the treatment of hospitalized patients with COVID-19: An individual participant data meta-analysis": S4 Fig

### S4 Fig. Posterior Predictive Check of Primary Outcomes by Study

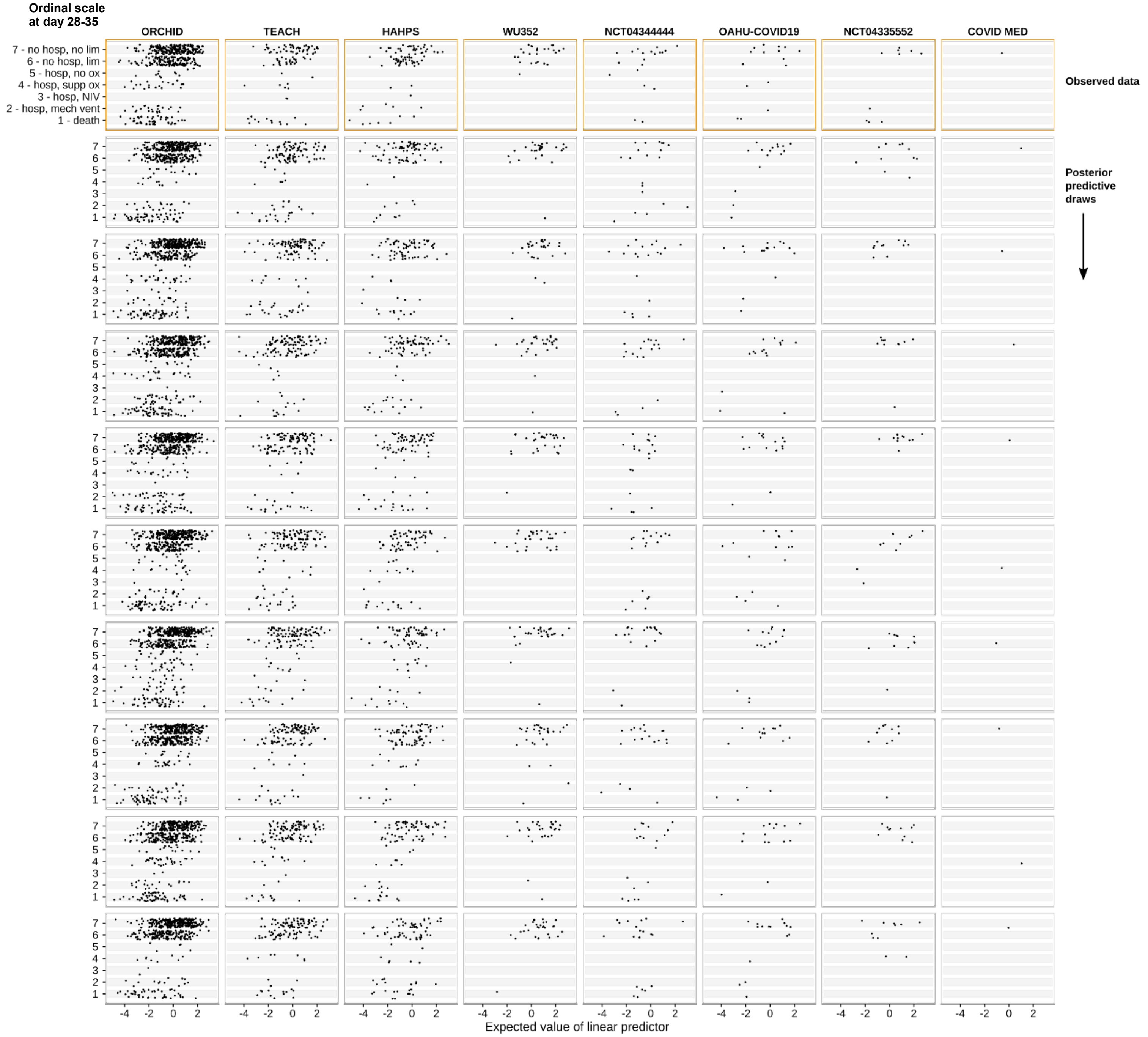


Posterior predictive check of the main analysis model. Shown are observed outcome data (first row) and draws for the posterior predictive distribution (subsequent rows) of the ordinal outcome scale at day 28-35, plotted against the expected linear predictor for each individual. Each column corresponds to one study in our analysis. Data points have been jittered for clarity.
