## Supplementary material for "Hydroxychloroquine/chloroquine for the treatment of hospitalized patients with COVID-19: An individual participant data meta-analysis": S4 Table

**S4 Table. Primary, Secondary, and Safety Outcomes**

| Trial | Primary outcome | Secondary outcomes | Safety outcomes |
|---|---|---|---|
| ORCHID | COVID Ordinal Outcomes Scale on Study Day 15 | • Time to recovery, defined as time to reaching level 5, 6, or 7 on the COVID Outcomes Scale, which is the time to the earlier of final liberation from supplemental oxygen or hospital discharge<br>• All-location, all-cause 14-day mortality (assessed on Study Day 15)<br>• All-location, all-cause 28-day mortality (assessed on Study Day 29)<br>• COVID Ordinal Outcomes Scale on Study Day 3<br>• COVID Ordinal Outcomes Scale on Study Day 8<br>• COVID Ordinal Outcomes Scale on Study Day 29<br>• Composite of death or receipt of ECMO through Day 28<br>• Oxygen-free days through Day 28<br>• Ventilator-free days through Day 28<br>• Vasopressor-free days through Day 28<br>• ICU-free days through Day 28<br>• Hospital-free days through Day 28 | • Seizure<br>• Atrial or ventricular arrhythmia<br>• Cardiac arrest<br>• Elevation in aspartate, aminotransferase, or alanine aminotransferase to twice the local upper limit of normal<br>• Acute pancreatitis<br>• Acute kidney injury<br>• Receipt of renal replacement therapy |
| WU352 | Time (hours) from randomization to recovery defined as (1) absence of fever, as defined as at least 48 hours since last temperature ≥38.0°C without the use of fever-reducing medications AND (2) absence of symptoms of greater than mild severity for 24 hours AND (3) not requiring supplemental oxygen beyond pre-COVID baseline AND (4) freedom from mechanical ventilation or death | • Time to resolution of fever defined as at least 48 hours since last temperature ≥38.0°C without the use of fever-reducing medications<br>• Time to improvement in symptoms (scored as mild or absent and remained so for 24 hour)<br>• Mean improvement in symptom from baseline<br>• Duration of hospitalization<br>• Proportion requiring supplementary oxygen (above baseline usage) at any time during follow-up<br>• Duration (days) of requirement for supplementary oxygen<br>• Proportion requiring ICU admission at any time during follow-up<br>• Proportion requiring mechanical ventilation at any time during follow-up<br>• Ventilator free days<br>• All-cause mortality | • Serious Adverse Events that occur within 6 weeks after randomization<br>• Unanticipated problem (UP)<br>• Unexpected adverse drug event (UADE) |
| NCT04335552 (Duke) | WHO ordinal scale measured at 14 days after enrollment | • Death during the index hospitalization<br>• Number of days on mechanical ventilation<br>• Proportion of patients not receiving mechanical ventilation at baseline who progress to requiring mechanical ventilation | • Arrhythmias (ventricular)<br>• Hepatic failure<br>• Bone marrow failure<br>• Aplastic anemia |

| Trial | Primary outcome | Secondary outcomes | Safety outcomes |
|---|---|---|---|
| | | during the index hospitalization<br>• WHO ordinal scale measured at 28 days after enrollment<br>• Hospital length of stay in days for the index hospitalization<br>• Days of fever (temperature ≥38.0) after randomization<br>• Days on supplemental oxygen after randomization<br>• All-cause study medication discontinuation<br>• Drug-associated adverse events of special interest | • Prolonged QT interval<br>• Angioedema<br>• Exfoliative dermatitis<br>• Acute generalized exanthematous pustulosis (AGEP)<br>• Psychosis<br>• Suicidal ideation<br>• Seizure<br>• SAEs that occur between initial dose of study medication and day 14 |
| TEACH | A severe disease progression composite endpoint defined by the occurrence of any of the following: mortality, ICU admission, invasive mechanical ventilation, ECMO, and/or hypotension requiring vasopressor support by the 14-day post-treatment evaluation | • Composite outcome (mortality, ICU admission, invasive mechanical ventilation, ECMO, and/or hypotension requiring vasopressor support) at 30 days<br>• Individual components of the composite endpoint (mortality, ICU admission, invasive mechanical ventilation, ECMO, and/or hypotension requiring vasopressor support) by EOT, PTE, and 30 days of treatment<br>• Hospital length of stay<br>• Days of fever<br>• Days of non-invasive ventilator use<br>• Days of non-rebreather mask oxygen supplementation<br>• Cytokine release syndrome grading scale<br>• Percentage of subjects reporting each severity score on 8-point ordinal scale D1 and EOT<br>• Percentage of subjects with QTc prolongation at EOT<br>• SARS-CoV-2 viral eradication from nasopharyngeal specimens at EOT, measured by RT-PCR<br>• Change from baseline AST, ALT, creatinine, glucose, white blood cell count, lymphocyte percentage, hemoglobin, platelets, total bilirubin, LDH, CRP, and IL-6 at EOT | • Primary Safety Composite: Cumulative incidence of SAEs through day 30, grade 3 or 4 AEs through day 30, and/or discontinuation of therapy (for any reason) |
| COVID MED | NIAID COVID-19 Ordinal Severity Score | • Mortality<br>• Hospital/ICU/ventilator length of stay (LOS)<br>• Reason for ventilator discontinuation (if applicable)<br>• Antibiotics use<br>• Symptoms-based COVID-19 severity score | • Complications<br>• AEs/SAEs |
| HAHPS | WHO COVID Ordinal Outcomes Scale at 14 days | • Hospital-free days at 28 days (calculated as a worst-rank ordinal)<br>• Ventilator-free days at 28 days (calculated as a worst-rank ordinal) | • Counts (proportions) of adverse events<br>• Those listed in the package insert for hydroxychloroquine and azithromycin<br>• QT interval |

| Trial | Primary outcome | Secondary outcomes | Safety outcomes |
|---|---|---|---|
| | | • ICU-free days at 28 days (calculated as a worst-rank ordinal)<br>• Time to a 1-point decrease in the WHO ordinal recovery score | • Arrythmia |
| NCT04344444 (University Medical Center New Orleans) | Composite incidence of death, transfer to ICU, initiation of mechanical ventilation, or initiation of ECMO | • Toxicities of the drugs<br>• SARS-CoV2 viral load over time<br>• Length of hospital stay<br>• Number of ICU days<br>• Rate of readmission after hospital discharge<br>• Duration of symptoms | • AEs monitored daily, including drug toxicity.<br>• QTc monitoring via telemetry |
| OAHU-COVID19 | Clinical status (on a 7-point ordinal scale) at day 15 | • Time to an improvement of one category from admission using an ordinal scale<br>• Subject clinical status using ordinal scale at days 3, 5, 8, 11, and 28<br>• Mean change in the ordinal scale from baseline to days 3, 5, 8, 11, 15, and 28 from baseline<br>• The time to discharge or to a National Early Warning Score (NEWS) of ≤2 and maintained for 24 hours, whichever occurs first<br>• Oxygenation free days in the first 28 days<br>• Incidence and duration of new oxygen use during the study<br>• Ventilator-free days in the first 28 days<br>• Incidence and duration of new mechanical ventilation use during hospitalization<br>• Duration of hospitalization (days) | • 28-day mortality<br>• Cumulative incidence of serious adverse events (SAEs) through 28-day follow-up<br>• Cumulative incidence of Grade 3 and 4 adverse events<br>• Discontinuation of hydroxychloroquine for any reason<br>• Changes in WBC, Hb, Plt, Creat, CrCl, Gluc, Tot Bili, ALT, AST over time |
