## Supplementary material for "Hydroxychloroquine/chloroquine for the treatment of hospitalized patients with COVID-19: An individual participant data meta-analysis": S5 Fig

### S5 Fig. Exploratory Analysis of Time Between Symptom Onset and Enrollment

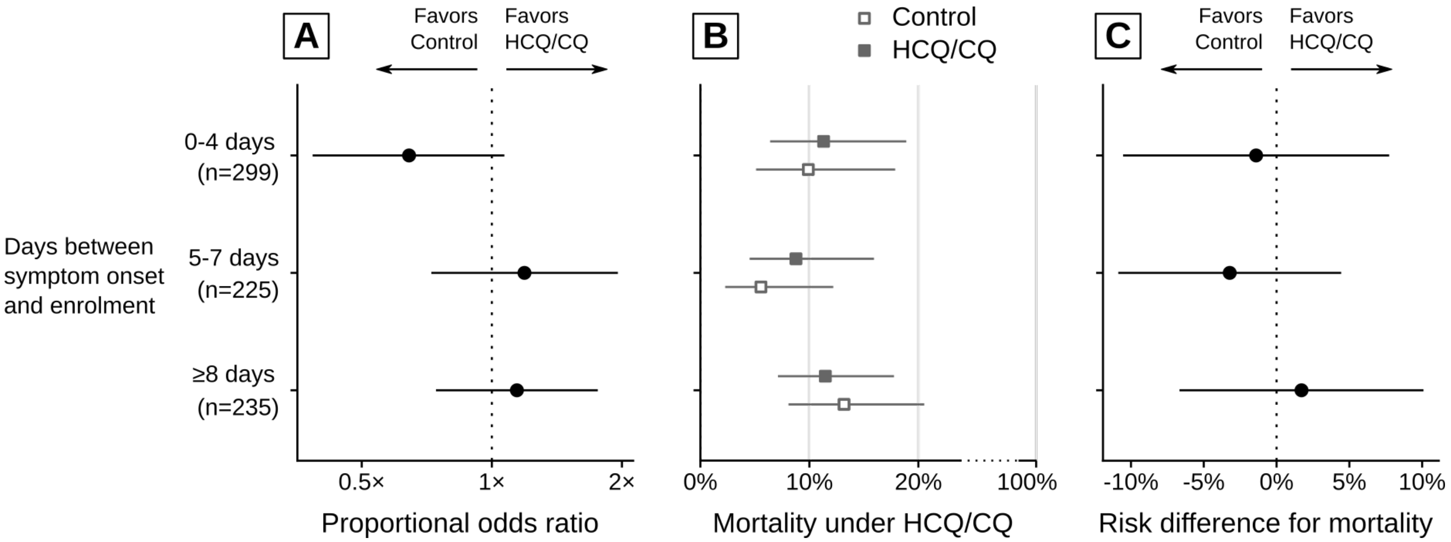


Post-hoc exploratory analysis of subgroups based on time between symptom onset and enrollment. Subgroups are based on approximate tertiles. Shown are (A) proportional odds ratios from models fit by maximum likelihood within each subgroup, with 95% CIs; (B) the empirical risk of survival for each treatment group within each subgroup, with 95% CIs; and (C) empirical risk differences within each subgroup, with 95% CIs.

HCQ/CQ indicates hydroxychloroquine or chloroquine.
