## Supplementary material for "Hydroxychloroquine/chloroquine for the treatment of hospitalized patients with COVID-19: An individual participant data meta-analysis": S5 Table

### S5 Table. Trial Characteristics: Treatment Groups, Participant Assessment, and Inclusion/Exclusion Criteria

| Trial (blinding) | Planned sample size | Age | Treatment groups | Treatment days | Dose by treatment group | Participant assessment | Inclusion Criteria | Exclusion Criteria |
|---|---|---|---|---|---|---|---|---|
| ORCHID<br><br>NCT04332991, Vanderbilt University, Massachusetts General, and PETAL Network<br><br>(Blinded) [1] | 510 hospitalized participants | ≥18 years | 2 | 5 days | **HCQ:** Days 1-2 400 mg twice daily; Days 3-10 200 mg twice daily;<br>**Placebo:** Matching placebo enterally twice daily matching the dosing regimen for HCQ | 12 months (Daily assessments Days 1-5, 8, 15, and 29, Months 3, 6, and 12) | • Age ≥18 years<br>• Currently hospitalized or in an emergency department with anticipated hospitalization.<br>• Symptoms of acute respiratory infection, defined as one or more of the following:<br>a. Cough<br>b. Fever (>37.5° C / 99.5° F)<br>c. Shortness of breath (operationalized as any of the following: subjective shortness of breath reported by patient or surrogate; tachypnea with respiratory rate ≥22 /minute; hypoxemia, defined as SpO2 <92% on room air, new receipt of supplemental oxygen to maintain SpO2 ≥92%, or increased supplemental oxygen to maintain SpO2 ≥92% for a patient on chronic oxygen therapy).<br>d. Sore throat<br>• Laboratory-confirmed SARS-CoV-2 infection within the past 10 days prior to randomization | • Prisoner<br>• Pregnancy<br>• Breast feeding<br>• Unable to randomize within 10 days after onset of acute respiratory infection symptoms<br>• Unable to randomize within 48 hours after hospital arrival<br>• Seizure disorder<br>• Porphyria cutanea tarda<br>• QTc >500 ms on electrocardiogram within 72 hours prior to enrollment<br>• Diagnosis of Long QT syndrome<br>• Known allergy to hydroxychloroquine, chloroquine, or amodiaquine<br>• Receipt in the 12 hours prior to enrollment, or planned administration during the 5-day study period that treating clinicians feel cannot be substituted for another medication, of any of the following:<br>◦ amiodarone; cimetidine; dofetilide; phenobarbital; phenytoin; sotalol<br>• Receipt of >1 dose of hydroxychloroquine or chloroquine in the 10 days prior to enrollment<br>• Inability to receive enteral medications<br>• Refusal or inability to be contacted on Day 15 for clinical outcome assessment if discharged prior to Day 15<br>• Previous enrollment in this trial<br>• The treating clinical team does not believe equipoise exists regarding the use of hydroxychloroquine for the treatment of this patient |
| WU352<br><br>NCT04341727, Washington University<br><br>(Open-label) | 500 non-ventilated hospitalized participants expected; 30 participants | ≥18 years | 4 | 5 days | **HCQ:** Day 1 400 mg twice daily; days 2-5 200 mg twice daily<br>**HCQ + AZM**: HCQ: Day 1 400 mg twice daily; Days 2- | 6 weeks (Daily assessments Days 1-14, Weeks 3, 4, 5, and 6) | • Hospitalization for management of SARS CoV-2 infection<br>• Positive SARS CoV-2 test<br>• Age ≥18 years<br>• Provision of informed consent | • Contraindication or allergy to chloroquine, hydroxychloroquine or azithromycin<br>• Current use hydroxychloroquine, chloroquine or azithromycin<br>• Concurrent use of another investigational agent<br>• Invasive mechanical ventilation |

| Trial (blinding) | Planned sample size | Age | Treatment groups | Treatment days | Dose by treatment group | Participant assessment | Inclusion Criteria | Exclusion Criteria |
|---|---|---|---|---|---|---|---|---|
| | when study closed | | | | 5 200 mg twice daily; AZM: Day 1 500 mg once, Days 2-5 250 mg once daily<br>**CQ**: Day 1 1,000 mg once, followed by 500 mg in 12 hours; Days 2-5 500 mg orally twice daily<br>**CQ + AZM**: CQ: Day 1 1000 mg once, followed by 500 mg in 12 hours; Days 2-5 500 mg orally twice daily; AZM: Day 1 500 mg once, Days 2-5 250 mg once daily | | • Electrocardiogram (ECG) ≤48 hours prior to enrollment<br>• Complete blood count, glucose-6 phosphate-dehydrogenase (G6PD), comprehensive metabolic panel and magnesium ≤48 hours prior to enrollment from standard of care<br>• If participating in sexual activity that could lead to pregnancy, individuals of reproductive potential who can become pregnant must agree to use contraception throughout the study. At least one of the following must be used throughout the study:<br>◦ Condom (male or female) with or without spermicide<br>◦ Diaphragm or cervical cap with spermicide<br>◦ Intrauterine device (IUD)<br>◦ Hormone-based contraceptive | • Participants who have any severe and/or uncontrolled medical conditions such as: unstable angina pectoris; symptomatic congestive heart failure; myocardial infarction; cardiac arrhythmias or known prolonged QTc >470 males, >480 female on ECG; pulmonary insufficiency; epilepsy (interaction with chloroquine)<br>• Prior retinal eye disease<br>• Concurrent malignancy requiring chemotherapy<br>• Known Chronic Kidney disease, eGFR<10 or dialysis<br>• G-6-PD deficiency, if unknown requires G6PD testing prior to enrollment<br>• Known Porphyria<br>• Known myasthenia gravis<br>• Currently pregnant or planning on getting pregnant while on study<br>• Breast feeding<br>• AST/ALT >five times the upper limit of normal ULN<br>• Bilirubin >five times the UL<br>• Magnesium <1.4 mEq/L<br>• Calcium <8.4mg/dL >10.6mg/dL<br>• Potassium <3.3 >5.5 mEg/L<br>• Current concomitant use of contraindicated drugs including antiarrhythmics, antidepressant, anticonvulsants |
| NCT04335552, Duke University<br><br>(Open-label) | 500 hospitalized participants | ≥12 years or older | 2 | 5 days | **Arm 1**: Supportive care alone<br>**Arm 2**: Supportive care + HCQ (Day 1 800 mg once; Days 2-5 600 mg once daily)<br>**Arm 3**: Supportive care + AZM (Day 1 500 mg once; Days 2-5 250 mg once daily)<br>**Arm 4**: Supportive care + HCQ (Day 1 | 45 days (Daily assessment Days 1-14, 28, and 45) | • Admitted to participating hospital with symptoms suggestive of SARS-CoV-2 infection OR develop symptoms of SARS-CoV-2 during hospitalization<br>• Subject (or legally authorized representative) can provide written informed consent (in English or Spanish) affirming intention to comply with planned study procedures prior to enrollment | • Participating in any other clinical trial of an experimental agent for SARS-CoV-2<br>• On hydroxychloroquine at any time during hospitalization, or within 180 days of hospitalization for COVID-19 regardless of indication<br>• History of cirrhosis, long QT syndrome or porphyria of any classification<br>• Most recent ECG prior to time of screening with QTc of ≥500 msec<br>• Known hypersensitivity to hydroxychloroquine or 4-aminoquinoline derivatives<br>• Weight less than 40 kg |

| Trial (blinding) | Planned sample size | Age | Treatment groups | Treatment days | Dose by treatment group | Participant assessment | Inclusion Criteria | Exclusion Criteria |
|---|---|---|---|---|---|---|---|---|
| | | | | | 800 mg once; Days 2-5 600 mg once daily) + AZM (Day 1 500 mg once; Days 2-5 250 mg once daily) | | • Male or female aged 12 years or older at the time of enrollment<br>• Has laboratory-confirmed SARS-CoV-2 infection determined by a validated nucleic acid amplification assay (public health or commercial) in any respiratory specimen collected within 14 days of randomization<br>• Illness of any duration that includes:<br>◦ Radiographic evidence of pulmonary infiltrates (chest X-ray or CT scan) OR<br>◦ Clinical documentation of lower respiratory symptoms (cough, shortness of breath, or wheezing) OR<br>◦ Any documented SpO2 ≤94% on room air OR<br>◦ Any inpatient initiation of supplemental oxygen regardless of documented cause | • Death anticipated within 48 hours of enrollment<br>• Inability to obtain informed consent from the patient or designated medical decision maker |
| TEACH<br><br>NCT04369742, New York University<br><br>(Blinded) [2] | 626 hospitalized adult and pediatric participants | >0 | 2 | 5 days | **HCQ:** Day 1 400 mg twice daily; Days 2-5 200 mg twice daily<br>**Placebo:** Day 1 calcium citrate 400 mg twice daily; Days 2-5 200 mg twice daily | 30 days (Baseline, End of treatment, Days 6, 14, and 30) | • Hospitalized with symptoms consistent with COVID-19 including but not limited to any of the following: fever (documented or subjective), cough, dyspnea, diarrhea, nausea, diffuse myalgias, and/or anosmia<br>• Informed consent signed by patient (if ≥18 years old) or parent (if <18 years old). Additionally, assent will be obtained from children ages 7 and older who are capable of providing assent. Adults who are unable to provide informed consent may be consented by legally authorized representative (see 13.3.3). | • Presence of the primary endpoint (ICU admission, mechanical ventilation, ECMO, and/or vasopressor requirement) at time of randomization.<br>• Treatment with CQ or CQ within the 30 days prior to the start of the study drug treatment.<br>• Unable to take oral medications.<br>• History of allergic reaction or intolerance to CQ or CQ.<br>• Baseline corrected QTc interval (>500 milliseconds, gender neutral) history of congenital QTc prolongation, and/or history of cardiac arrest.<br>• Concomitant therapy with flecainide, amiodarone, digoxin, procainamide, propafenone, thioridazine, or pimozide<br>• History of retinal disease including a documented history of diabetic retinopathy. |

| Trial (blinding) | Planned sample size | Age | Treatment groups | Treatment days | Dose by treatment group | Participant assessment | Inclusion Criteria | Exclusion Criteria |
|---|---|---|---|---|---|---|---|---|
| | | | | | | | • Positive SARS-CoV-2 RT-PCR testing (nasopharyngeal, oropharyngeal, sputum and/or bronchoalveolar lavage) The testing may:<br>◦ Occur up to ≤72h prior to informed consent of participation in the study<br>◦ Be undertaken either on-site or in an external laboratory certified by New York State to run testing for SARS-CoV-2 | • Known history of G6PD deficiency.<br>Pediatric Exclusion Criteria:<br>• Baseline QTc >470 ms in males, >480 ms in females (post puberty) or QTc >460 ms in males, >470 ms in females (pre puberty)<br>• History of congenital QT prolongation (LQTS) and/or history of cardiac arrest.<br>• Family history of LQTS<br>• Presence of Concomitant therapy QT prolongation: Medications will be checked against a list on www.CredibleMeds.com, and those on concomitant medications with significant QT-prolonging potential will be excluded<br>• Basic metabolic panel (BMP) not performed within 72 hours of enrollment<br>• Presence of uncorrected hypokalemia (<3.4 mmol/L), hypocalcemia (<9.0 mg/dL, and/or hypomagnesemia (<1.7 mg/dL) on most recent BMP (within 72 hours of enrollment). |
| COVID MED<br><br>NCT04328012, Bassett Medical Center<br><br>(Blinded) | 4,000 hospitalized participants | ≥18 years | 4 | Up to 14 days | **Arm 1**: standard care and lopinavir/ritonavir: Dosing: 400 mg/100 mg twice daily for 5-14 days<br>**Arm 2**: standard care and **HCQ**: Day 1 400 mg twice daily; Days 2-14 200 mg twice daily<br>**Arm 3**: standard care and losartan<br>•Losartan 25 mg once daily for 5-14 days<br>•Placebo (Tic Tacs in blank capsules) once daily for 5-14 days to replicate/control for 'bid dosing'<br>**Arm 4**: standard | 60 days (Baseline, Day 1-7, Day 14, Day 30, Day 60) | • Hospitalized patient<br>• Age ≥18 years<br>• Able to ingest oral medication or be administered medication via gastric tube or equivalent<br>• Laboratory confirmation of SARS-CoV-2 infection within 1 week prior to randomization<br>• Randomization within 72 hr of hospital admission<br>• Negative pregnancy test for reproductive age women<br>• Patient or LAR able to provide informed consent | General (all groups) exclusions:<br>• End stage renal disease (ESRD) NOT undergoing renal replacement therapy<br>• Severe hepatic insufficiency (LFTs >5 times the upper limit of normal or known ESLD or cirrhosis)<br>• Nausea/vomiting or aspiration risk precluding oral medications unless can be given by gastric tube<br>• Use of another SARS-CoV-2 directed medication empirically or within another clinical trial within the prior week<br>• Pregnancy or breast feeding<br>• Absence of dependable contraception in reproductive age women<br>• Inability to obtain or declined informed consent<br><br>Hydroxychloroquine group exclusions:<br>• Allergy or intolerance to HCQ (or CQ)<br>• Already taking HCQ or CQ (within 1 month) |

| Trial (blinding) | Planned sample size | Age | Treatment groups | Treatment days | Dose by treatment group | Participant assessment | Inclusion Criteria | Exclusion Criteria |
|---|---|---|---|---|---|---|---|---|
| | | | | | care and **placebo**: Placebo (Tic Tacs in blank capsules) twice daily for 5-14 days | | | • Recent malaria exposure (within 1 month)<br>• History or current cardiac diseases (heart failure, ventricular arrhythmias, LBBB or RBBB, QTc prolongation)<br>• History of retinopathy<br>• Severe hypoglycemia<br>• Auditory disorders<br>• Known G6PD deficiency<br>• Porphyria or psoriasis<br>• Severe active alcohol use disorder<br>• Seizure disorder<br>• Co-administration of hepatotoxic agents<br>• Co-administration with certain drugs due to CYP3A interactions if taken in <24 hr |
| HAHPS<br><br>NCT04329832, Intermountain Health Care<br><br>(Open-label) [3,4] | 300 hospitalized participants | ≥18 years | 2 | 5 days | **HCQ:** Day 1 400 mg twice daily; Days 2-5 200 mg twice daily<br>**AZM**: Day 1 500 mg once; Days 2-5 250 mg once daily | 6 months (Daily assessments Days 1-7 and 14, Month 6) | • Adult (age ≥18 years)<br>• Confirmed OR suspected COVID-19<br>Confirmed: Positive assay for COVID-19 within the last 10 days<br>Suspected: Pending assay for COVID-19 WITH high clinical suspicion<br>• Scheduled for admission or already admitted to an inpatient bed | • Allergy to hydroxychloroquine or azithromycin<br>• History of bone marrow transplant<br>• Known G6PD deficiency<br>• Chronic hemodialysis or glomerular filtration rate <20 ml/min<br>• Psoriasis<br>• Porphyria<br>• Concomitant use of digitalis, flecainide, amiodarone, procainamide, propafenone, cimetidine, dofetilide, phenobarbital, phenytoin, or sotalol<br>• Known history of long QT syndrome<br>• Current known QTc >500 msec<br>• Pregnant or nursing<br>• Prisoner<br>• Weight <35 kg<br>• Seizure disorder<br>• Severe liver disease<br>• Outpatient use of hydroxychloroquine for treatment of a disease other than COVID-19 OR has received more than 2 days of hydroxychloroquine or azithromycin for suspected or confirmed COVID-19<br>• Patient has recovered from COVID-19 and/or is being discharged from the hospital on day of enrollment<br>• Treating physician refuses to allow patient participation in the study |

| Trial (blinding) | Planned sample size | Age | Treatment groups | Treatment days | Dose by treatment group | Participant assessment | Inclusion Criteria | Exclusion Criteria |
|---|---|---|---|---|---|---|---|---|
| | | | | | | | | • Unable to obtain informed consent<br>• Prior enrollment in this study |
| NCT04344444, University Medical Center New Orleans<br><br>(Open-label) | 600 hospitalized participants | ≥18 years | 3 | 5 days | **Control**: Supportive Care Only<br>**HCQ:** Day 1 400 mg twice daily; Days 2-5 200 mg twice daily<br>**HCQ + AZM**: HCQ: Day 1 400 mg twice daily; Days 2-5 200 mg twice daily; AZM: Day 1 500 mg once; Days 2-5 250 mg daily | 30 days (Daily assessments Days 1-14, and 30) | • Age greater than 18 years<br>• Positive SARS-CoV-2 testing or consistent clinical syndrome (based on clinical picture e.g., characteristic infiltrates on chest x-ray, laboratory findings, and with agreement by two physicians) in patients under investigation (PUIs).<br>• Oxygen saturation of >94% on room air with defined risk factors consistent with moderate disease OR oxygen saturation of <94% on room air consistent with severe disease<br>• Ability and willingness to comply with study procedures | • QTc greater than 450 milliseconds on screening EKG or telemetry<br>• Pregnant or lactating women<br>• Inability to take oral pills or inability to use a feeding tube<br>• Inability to obtain informed consent either from the patient or from the next of kin if patient is incapacitated. For the purpose of this study obtaining a verbal consent from a family member on the phone with a witness will be considered acceptable since there is a "no visitor" policy in force at hospitals.<br>• Patients requiring ICU level care<br>• Use of azithromycin or hydroxychloroquine within 30 days prior to admission |
| OAHU-COVID19<br><br>NCT04345692, Queen's Medical Center -Honolulu<br><br>(Open-label) | 350 hospitalized participants | 18-95 years | 2 | 5 days | **Usual Care + HCQ:** Day 1 400 mg twice daily; Days 2-5 200 mg twice daily<br>**Usual care** | BL, Days 3, 5, 8, 11, and 28 | | |
