## Supplementary material for "Hydroxychloroquine/chloroquine for the treatment of hospitalized patients with COVID-19: An individual participant data meta-analysis": S6 Table

### S6 Table. Merging Trial Arms

| Trial | Treatment groups | Treatment days | Treatment or control? | Dose by treatment group |
|---|---|---|---|---|
| ORCHID | 2 | 5 days | T | HCQ: Day 1 400 mg twice daily; Days 2-5 200 mg twice daily |
| | | | C | Placebo |
| WU352 | 4 | 5 days | T | HCQ: Day 1 400 mg twice daily; Days 2-5 200 mg twice daily |
| | | | T | HCQ + AZM: HCQ: Day 1 400 mg twice daily; Days 2-5 200 mg twice daily; AZM: Day 1 500 mg once; Days 2-5 250 mg once daily |
| | | | T | CQ: Day 1 1000 mg once followed by 500 mg in 12 hours; Days 2-5 500 mg twice daily |
| | | | T | CQ + AZM: CQ: Day 1 1000 mg once followed by 500 mg in 12 hours; Days 2-5 500 mg twice daily; AZM: Day 1 500 mg once; Days 2-5 250 mg once daily |
| NCT04335552 (Duke University) | 2 | 5 days | C | Arm 1: Supportive care alone |
| | | | T | Arm 2: Supportive care plus HCQ (Day 1 800 mg once; Days 2-5 600 mg once daily) |
| | | | C | Arm 3: Supportive care plus azithromycin (Day 1 500 mg once; Days 2-5 250 mg once daily) |
| | | | T | Arm 4: Supportive care plus HCQ (Day 1 800 mg once; Days 2-5 600 mg once daily) plus azithromycin (Day 1 500 mg once; Days 2-5 250 mg once daily) |
| TEACH | 2 | 5 days | T | HCQ: Day 1 400 mg twice daily; Days 2-5 200 mg twice daily |
| | | | C | Placebo |
| COVID MED | 4 (only Arms 2 and 4 included herein) | 14 days | C | Arm 1: Standard care and lopinavir/ritonavir: Dosing: 400 mg/100 mg twice daily for 5-14 days |
| | | | T | Arm 2: Standard care and HCQ: Day 1 400 mg twice daily; Days 2-5 200 mg twice daily |
| | | | C | Arm 3: Standard care and losartan: Losartan 25 mg daily for 5-14 days; placebo daily for 5-14 days to replicate/control for twice-daily dosing |
| | | | C | Arm 4: Standard care and placebo: Placebo twice daily for 5-14 days |
| HAHPS | 2 | 5 days | T | HCQ: Day 1 400 mg twice daily; Days 2-5 200 mg twice daily |
| | | | C | AZM: Day 1 500 mg once; Days 2-5 250 mg once daily |
| NCT04344444 (University Medical Center New Orleans) | 3 | 5 days | C | Control: Supportive Care Only |
| | | | T | HCQ: Day 1 400 mg twice daily; Days 2-5 200 mg twice daily |
| | | | T | HCQ + AZM: HCQ: Day 1 400 mg twice daily; Days 2-5 200 mg twice daily; AZM: Day 1 500 mg once; Days 2-5 250 mg once daily |
| OAHU-COVID19 | 2 | 5 days | T | HCQ: Day 1 400 mg twice daily; Days 2-5 200 mg twice daily |
| | | | C | Usual care |
